# Evaluation of New and Repurposed Tools to Assess Post-Tuberculosis Lung Disease in Adolescents: A Cross-Sectional Analysis

**DOI:** 10.1101/2025.06.29.25330520

**Authors:** Joshua Ray Tanzer, Leonid Lecca, Betsabe Roman Sinche, Victoria Sanchez Guzman, Justin J. Wyda, Anthony L. Byrne, Silvia S. Chiang

**Author notes:** Joint senior authors (Anthony L. Byrne, Silvia S. Chiang).

## Abstract

**Background:** Tuberculosis, even when successfully treated, frequently leads to long-term sequelae, known as post-tuberculosis lung disease (PTLD). Adolescents account for over 1 million incident tuberculosis cases each year, and PTLD in this population may contribute greatly to the global burden of chronic lung disease. However, research to better understand and prevent adolescent PTLD is hampered by uncertainty regarding which tools best assess respiratory disability and lung function in this population.

**Methods:** In this cross-sectional analysis of 101 adolescent tuberculosis survivors in Lima, Peru, we administered the St. George’s Respiratory Questionnaire (SGRQ) to assess respiratory disability, and spirometry and oscillometry to measure lung function. We used factor analysis, correlations, and structural equation modeling to assess reliability and validity of an abbreviated SGRQ, oscillometry, and spirometry.

**Results:** Our abbreviated, 18-item SGRQ had high reliability (omega ≥0.90), internal structure validity (factor loadings for most questions >0.75), and external validity (correlation: -0.62 with overall health rating). More participants were able to complete oscillometry vs. spirometry (100% vs. 91.1%, *p* <0.0001). Oscillometry metrics had higher reliability (0.82-0.88) than spirometry metrics (0.62-0.76). SGRQ scores had small correlations with spirometry and minimal correlations with oscillometry. The combination of the SGRQ, oscillometry, and spirometry demonstrated good model fit as an overall assessment of lung health.

**Conclusion:** Our findings support the combination of an abbreviated version of the SGRQ, spirometry, and oscillometry for evaluating adolescent PTLD. These results pave the way for critical research to better understand the long-term impacts of tuberculosis on adolescent lung health.

## INTRODUCTION

Globally, over 1 million adolescents (people 10-19 years old) fall sick with tuberculosis (TB) each year.^1^ Approximately 15.5 million people who had TB disease as adolescents (“adolescent TB survivors”) in the last 40 years are alive today.^2^ Even when successfully treated, pulmonary TB can lead to post-TB lung disease (PTLD), defined as respiratory disability – persistent symptoms and/or activity limitations – with impaired lung function.^3,4^ PTLD has been reported in 57-67% of adolescent TB survivors.^5–7^ Taken together with the high incidence of pulmonary TB in adolescents and the expected long duration of life ahead of them, these findings suggests that adolescent PTLD may account for a substantial burden of chronic lung disease globally.

Further research is needed to better characterize PTLD in adolescent TB survivors but is impeded by challenges to measuring both respiratory disability and lung function impairment. No questionnaires have been validated for PTLD-associated respiratory disability. Some PTLD studies have used the St. George’s Respiratory Questionnaire (SGRQ), which was originally developed for adults with chronic pulmonary disorders and has since been used for a wide range of respiratory diseases.^5,8–10^ However, the questionnaire is extensive, some items may be irrelevant to adolescents, and scoring is complicated.

Lung function impairment is determined using pulmonary function tests (PFTs), most commonly spirometry, which measures inspiratory and expiratory volume and speed. Although spirometry is relatively affordable and frequently used, it relies on correctly performed forced expiratory maneuvers, requiring the participant to exert substantial physical effort and the operator to correctly instruct and motivate the participant. Oscillometry, in contrast, is less commonly used but is much easier because it is performed during normal tidal breathing. Oscillometry utilizes vibrating sound waves to analyze pressure and flow of the respiratory system. It can better detect small airways disease compared to spirometry and uniquely measures lung tissue compliance to detect airway dysfunction or parenchymal damage.^11–13^ However, its feasibility and reliability have not been evaluated for TB survivors. Moreover, reference ranges are based on fewer than 400 participants from high-income settings, raising the question of whether they are appropriate for adolescents in low- and middle-income countries (LMICs), where TB is most prevalent.^14^

We conducted this study to validate and assess the performance of new and repurposed tools for adolescent PTLD in a LMIC setting. Specifically, we aimed to evaluate the (1) reliability and validity of an abbreviated, easier-to-score version of the SGRQ; (2) reference standards for oscillometry; (3) feasibility and reliability of oscillometry and spirometry; and (4) internal consistency and model fit of the overall assessment of lung health using all three tools.

## METHODS

### Study design and setting

This cross-sectional analysis used data from a prospective cohort study, Post-TB Teens, which we conducted from March 2022-September 2023 in Lima, Peru. The TB incidence in Peru is an estimated 173 per 100,000 per year.^15^ The primary aim of Post-TB Teens was to compare lung function and respiratory disability between adolescent TB survivors and healthy adolescents with no history of TB disease (“controls”). This current analysis sets the stage for the comparison by assessing the validity and reliability of the tools used to measure PTLD.

### Participant recruitment and eligibility

We identified TB survivors from Predict Teen, our previously completed study of adolescents who received treatment for rifampicin-susceptible TB at a health center run by the Ministry of Health.^16^ Study personnel recruited eligible participants in sequential order of their enrollment into Predict Teen. We included adolescents with pulmonary TB, with or without pleural effusion, and a treatment outcome of cure or treatment completion, as per World Health Organization reporting guidelines.^17^ Each TB survivor was matched 1:1 with a “control” of the same sex, age (within two years), and jurisdiction of the health center or the adjacent one. To identify controls, we screened household contacts of people with TB and the friends and family members of the enrolled TB survivors. The following exclusion criteria were applied to both groups: relapsed TB, use of glucocorticoids or other immunosuppressive agents within the past six months, pneumothorax or another lung lesion within the last six weeks, or surgery of the upper torso within the last four weeks.

### Data collection

TB survivors were evaluated at least four months after the end of TB treatment. Controls were evaluated as close as possible in time to their matched TB survivor. Assessments were rescheduled if the participant had signs/symptoms of a respiratory illness on the scheduled day of the assessment. Participants with TB symptoms (one or more of the following symptoms for at least two weeks: cough, fever, night sweats, weight loss, poor appetite, and persistent fatigue) were evaluated for TB. Those diagnosed with TB during the study were withdrawn, along with their matched pair.

Each lung health evaluation consisted of the following sequence of procedures: measurement of weight and height, pre-bronchodilator oscillometry (TremoFlo Airwave Oscillometer, Thorasys, Montreal, Canada),^18^ pre-bronchodilator spirometry (Easy-One Spirometer, NDD, Andover, USA),^19,20^ administration of salbutamol 200 mcg-400 mcg through a metered dose inhaler, the SGRQ in Spanish,^8^ post-bronchodilator oscillometry, and post-bronchodilator spirometry. Post-bronchodilator PFTs were performed at least 15 minutes after salbutamol administration.

Oscillometry and spirometry procedures and interpretation followed standards published by the American Thoracic Society and European Respiratory Society.^18^ Participants repeated the spirometry procedure until three suitable (quality score A-C) measurements were recorded.^19,20^ If participants were unable to complete at least one spirometry maneuver with a quality score of a C or better, the procedure was repeated as many times as tolerated in the same visit or at a second visit, depending on the participant’s willingness and availability. For spirometry, we recorded forced expiratory volume in one second (FEV1), functional vital capacity (FVC), and FEV1/FVC. For oscillometry, we recorded total airway resistance (R5) and reactance area (AX), also known as lung compliance. Additionally, we collected information on prior asthma diagnosis, tobacco use, and exposure to indoor biomass.

### Ethics

The institutional review boards of *Socios En Salud Sucursal Perú* and Brown University Health approved this study. Written informed consent was obtained from participants 18 years or older and parents/legal guardians of minors. Informed assent was obtained from minor participants.

### Analysis

We restricted most analyses to data from TB survivors, as they represent the target population for these evaluations. The exception was evaluation of oscillometry reference standards, for which we used data from healthy study participants with no history of TB disease. We conducted analyses using R (R Foundation for Statistical Computing, Vienna, Austria). Inference that a statistic is meaningful (i.e., *p* < 0.05) was based on 400 bootstrap confidence intervals. To address missing data, we used full information maximum likelihood estimation. We report confidence intervals for descriptive statistics and p-values for hypothesis testing (i.e., testing correlations or comparing proportions).

### Revision of the SGRQ

The SGRQ contains three subscales – symptoms, activities, and impact on daily life – and yields four scores: one for each subscale and an overall score. We removed questions from the SGRQ if >90% of participants selected the same answer choice, as items are useful only when there is variation in individual responses. However, we elected to keep questions that, despite meeting criteria for removal, were thought to be clinically important.

We simplified scoring by scaling each question from 0 to 1. A binary response of “true” is scored 1; “false,” 0. For multiple-choice questions with responses on a continuum (i.e., frequency or severity of symptoms), the responses are scaled between 0 and 1. The average for individual subsections or across all questions are calculated and multiplied by 100, resulting in scores that range from 0, indicating minimal impact of lung health on well-being, to 100, reflecting significant impact of lung health on well-being.

### Reliability and validity of the revised SGRQ

We applied factor analysis to evaluate the (1) internal consistency reliability and (2) internal structure validity of (a) each subscale (symptoms, activities, impact on daily life) and (b) the revised SGRQ in its entirety. We used coefficient omega to estimate internal consistency reliability, which indicates whether individual items within each subscale – or each subscale within the SGRQ – are related, thereby suggesting that they measure the same construct. Internal structure validity, measured through the calculation and comparison of factor loadings, indicates whether individual items contribute to the subscale – or whether individual subscales contribute to the overall SGRQ. To assess concurrent validity of the revised SGRQ, we evaluated correlations between each adolescent’s score and their rating of their overall health, scored as 1 (“very bad”), 2 (“bad”), 3 (“okay”), 4 (“good”), or 5 (“very good”).

In our setting and others, gender is associated with different concerns related to respiratory health; for instance, female gender is associated with less physical activity.^21^ Therefore, we stratified estimates of reliability and validity by gender.

### Oscillometry reference standards

To evaluate reference standards for oscillometry, we transformed R5 and AX for the controls into z-scores using the Oosteveen reference equations, which are based on 368 adults 18 years or older across Europe and Australia and incorporate height, weight, age, and sex. We plotted histograms of z-scores and visually compared the observed distributions against a normal distribution, which would represent a perfect fit of the reference equations to our population.

### Feasibility and reliability of oscillometry

We used the binomial test to compare the proportions of adolescents who successfully completed oscillometry vs. spirometry. To evaluate the internal consistency reliability of spirometry and oscillometry metrics, we applied latent variable structural equation modeling. Introducing latent variables helped account for the potential impacts of bronchodilation, as well as practice and fatigue, on lung function testing. The potential impacts of practice and fatigue are particularly important for spirometry, as a participant with good lung function may perform the challenging spirometry maneuver better at the second occasion, while a participant with poor lung function that is bronchodilator-unresponsive may perform worse due to the exhaustion. Thus, we specified latent variables for each pair of pre- and post-bronchodilation metrics to isolate the common aspects of breathing quality.^22^

### Reliability and model fit of the overall assessment of lung health

Finally, we examined the combination of SGRQ, spirometry, and oscillometry as an overall assessment of lung health. To assess internal consistency within this construct, we estimated correlations between latent variable scores of spirometry and oscillometry metrics, as well as revised and original SGRQ scores. Structural equation modeling model fit was examined to understand how well these combined measures represent lung health. A hierarchical latent variable representing the three SGRQ subscales was added to the structural equation model described above. Factor loadings were used to estimate reliability; covariances between latent variable residuals were specified in the model to improve estimation of underlying relationships unique to each trait as separate from relationships across traits. Latent variable weights and model fit statistics were estimated twice, using first the original and then the revised SGRQ. We assessed how well the specified model represented the data compared to a null model – which assumes no associations between any variables – using comparative fit index (CFI; acceptable values >0.90), root mean square error of approximation (RMSEA; acceptable values <0.10), and standardized root mean square residual (SRMR; acceptable values <0.10). Additionally, the chi-square statistic represented the model fit to data, with smaller or less significant values representing greater concordance between specified model and observed data.^23^

## RESULTS

We enrolled 102 TB survivors and 102 controls (Figure 1). One control developed TB disease during the study; therefore, he and his matched TB survivor were excluded from the analysis, leaving 101 participants in each group. At enrollment, the median ages and interquartile ranges (IQR) of TB survivors and healthy controls were 17 (16, 19) and 18 (17, 20) years, respectively. TB survivors were evaluated a median of 381 (IQR: 246, 517; range: 126-690) days after treatment completion. In both groups, 56 (55.4%) participants were male; 6 (5.9%) had prior asthma diagnoses; and none had HIV. Participants in both groups were minimally exposed to tobacco or indoor biomass; the median number of days of exposure was 0 (IQR: 0, 0) for both tobacco and indoor biomass, and the maximums were 1 and 0.25 days per week, respectively.

**Fig. 1:**
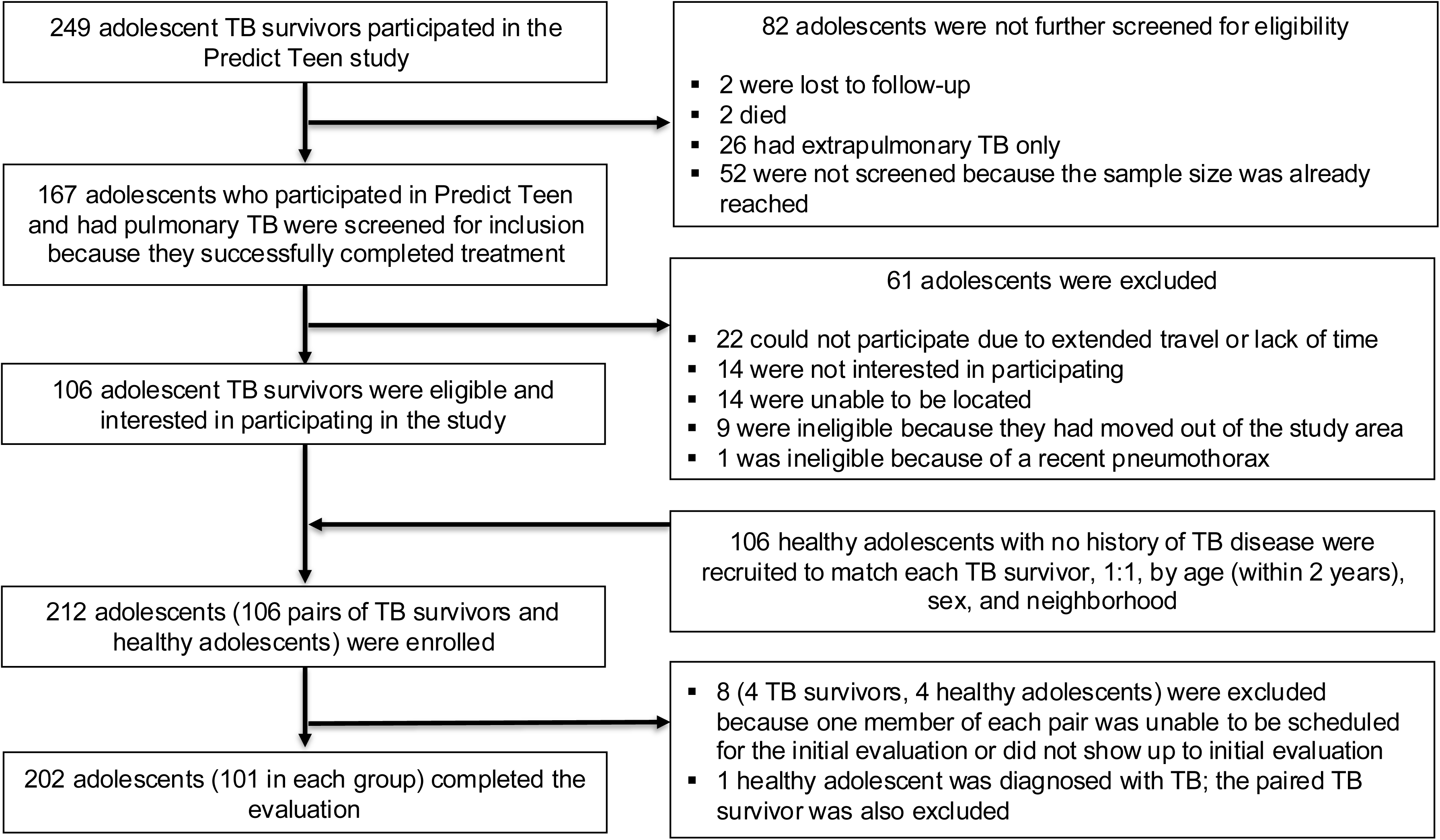
Participant screening, recruitment, enrollment, and retention.

The only missing variables were spirometry metrics in nine adolescents who could not perform the maneuver to acceptable technical standards.

### Revised SGRQ

The revised SGRQ included 18 questions, of which three met criteria for removal but were retained due to their clinical importance (Table 1; Appendix). The median score was 8.89 (IQR: 5.56, 20.87; range: 3.72-66.67). Median overall health rating was 4 (IQR: 4, 4; range: 2-5).

**Table 1:**
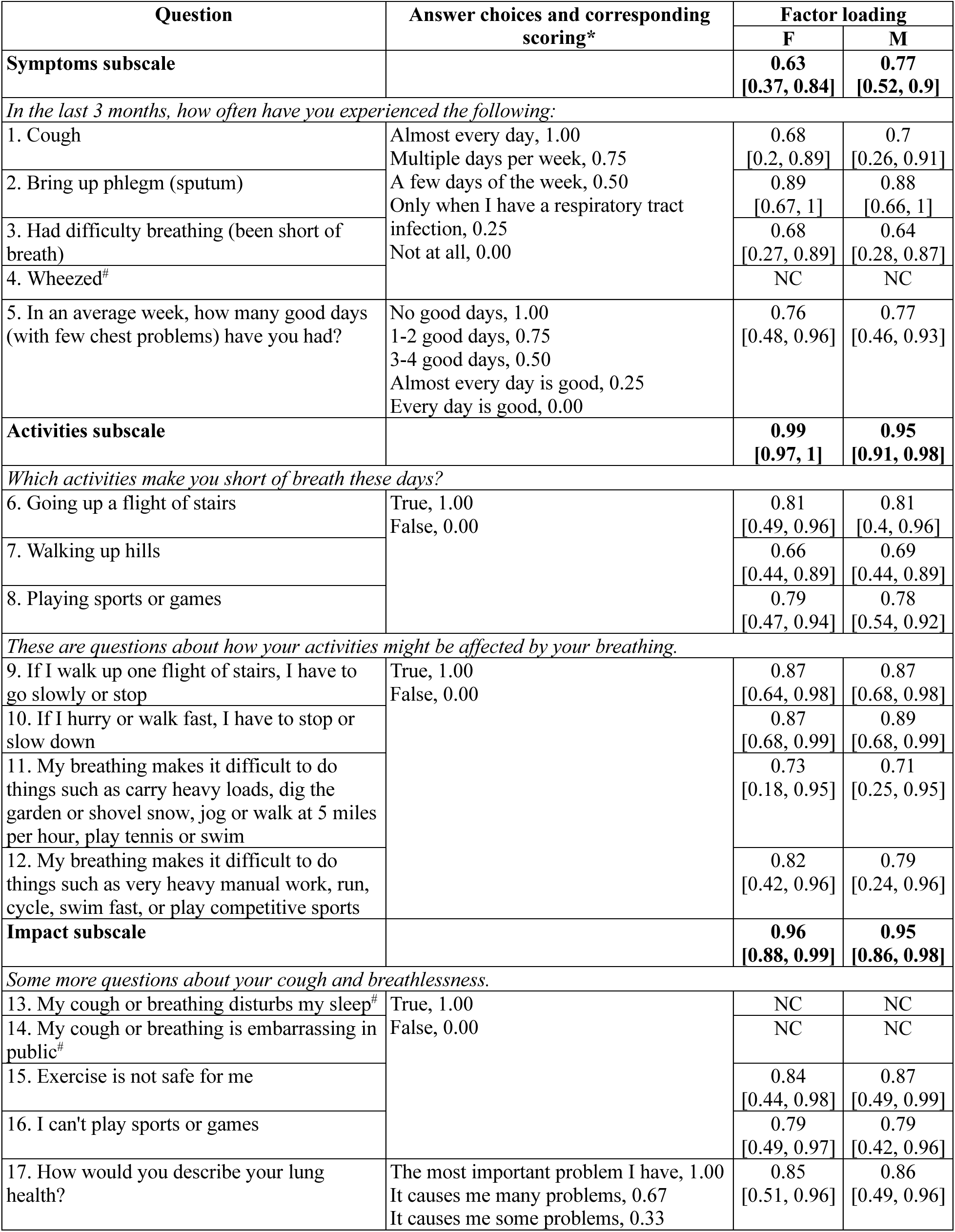
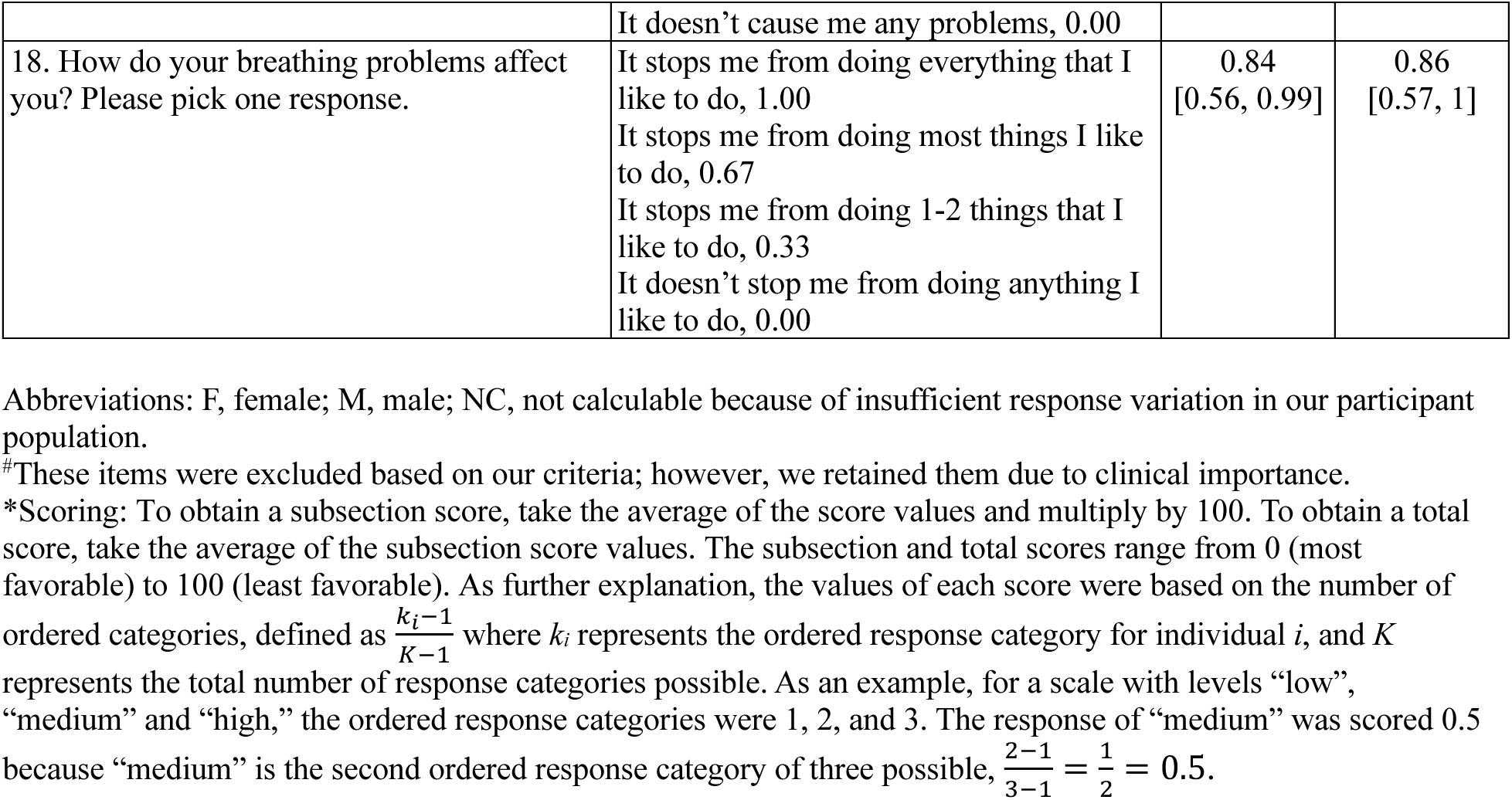
Revised St. George’s Respiratory Questionnaire: Retained Items, Scoring, and Factor Loadings.

Coefficient omega values of the symptom, activities, and impact subscales were 0.82 (95% CI: 0.72, 0.91), 0.92 (0.83, 0.95), and 0.89 (0.87, 0.96), respectively, for both genders. Coefficient omega for the entire SGRQ was 0.90 (95% CI: 0.84, 0.95) for females and 0.92 (95% CI: 0.86, 0.96) for males. Table 1 reports factor loadings of each individual item in relation to its subscale, as well as each of the three subscales in relation to the SGRQ in its entirety. Questions showed large positive associations, with most factor loadings exceeding 0.75; no gender differences were observed. Concurrent validity was -0.62 (95% CI: -0.81, -0.32) for both genders.

### Oscillometry and spirometry

All participants completed oscillometry. Nine of 101 (8.9%) TB survivors were unable to achieve a satisfactory spirometry maneuver (*p* < 0.0001). Distribution of R5 and AX values, after transformation into z-scores using the Oosteveen reference equations, approximated the normal distribution (Figure 2). Internal consistency reliability was high for oscillometry metrics, R5 and AX (Table 2). For spirometry, the only metric with acceptable reliability was FVC.

**Table 2:** Internal consistency reliability and correlations between St. George Respiratory Questionnaire scores, oscillometry metrics, and spirometry metrics.

| Row | Measure | 1 | 2 | 3 | 4 | 5 | 6 | 7 |
| --- | --- | --- | --- | --- | --- | --- | --- | --- |
| 1 | Revised SGRQ | **0.97**<br>[0.94, 0.99] |  |  |  |  |  |  |
| 2 | Original SGRQ | 0.95<br>($p < 0.001$)* | **0.99**<br>[0.95, 0.99] | | | | | |
| 3 | R5 | 0.02<br>($p = 0.79$) | -0.01<br>($p = 0.90$) | **0.88**<br>[0.81, 0.94] | | | | |
| 4 | AX | -0.07<br>($p = 0.48$) | -0.03<br>($p = 0.80$) | 0.49<br>($p < 0.001$)* | **0.82**<br>[0.73, 0.88] | | | |
| 5 | FEV<sub>1</sub> | 0.18<br>($p = 0.08$) | 0.15<br>($p = 0.16$) | -0.31<br>($p = 0.009$)* | -0.38<br>($p = 0.002$)* | **0.63**<br>[0.52, 0.70] | | |
| 6 | FVC | -0.05<br>($p = 0.58$) | -0.06<br>($p = 0.40$) | -0.11<br>($p = 0.43$) | -0.12<br>($p = 0.30$) | 0.38<br>($p < 0.001$)* | **0.76**<br>[0.71, 0.86] | |
| 7 | FEV<sub>1</sub>/FVC | 0.19<br>($p = 0.08$) | 0.18<br>($p = 0.10$) | -0.32<br>($p = 0.003$)* | -0.32<br>($p = 0.008$)* | 0.86<br>($p < 0.001$)* | -0.10<br>($p = 0.36$) | **0.62**<br>[0.53, 0.71] |

Estimates represent the correlations between latent variable scores. Bolded values along the diagonal represent omega coefficient estimates of reliability (with 95% confidence intervals in brackets) implied by the latent variable model. The research standard for acceptable reliability is 0.70. We did not perform hypothesis testing in the assessment of reliability and, thus, did not report $p$ -values. In contrast, $p$ -values are reported for non-bolded values in the off-diagonal cells to imply hypothesis testing against the null hypothesis of no association.
\* $p < 0.05$

**Fig. 2a:**
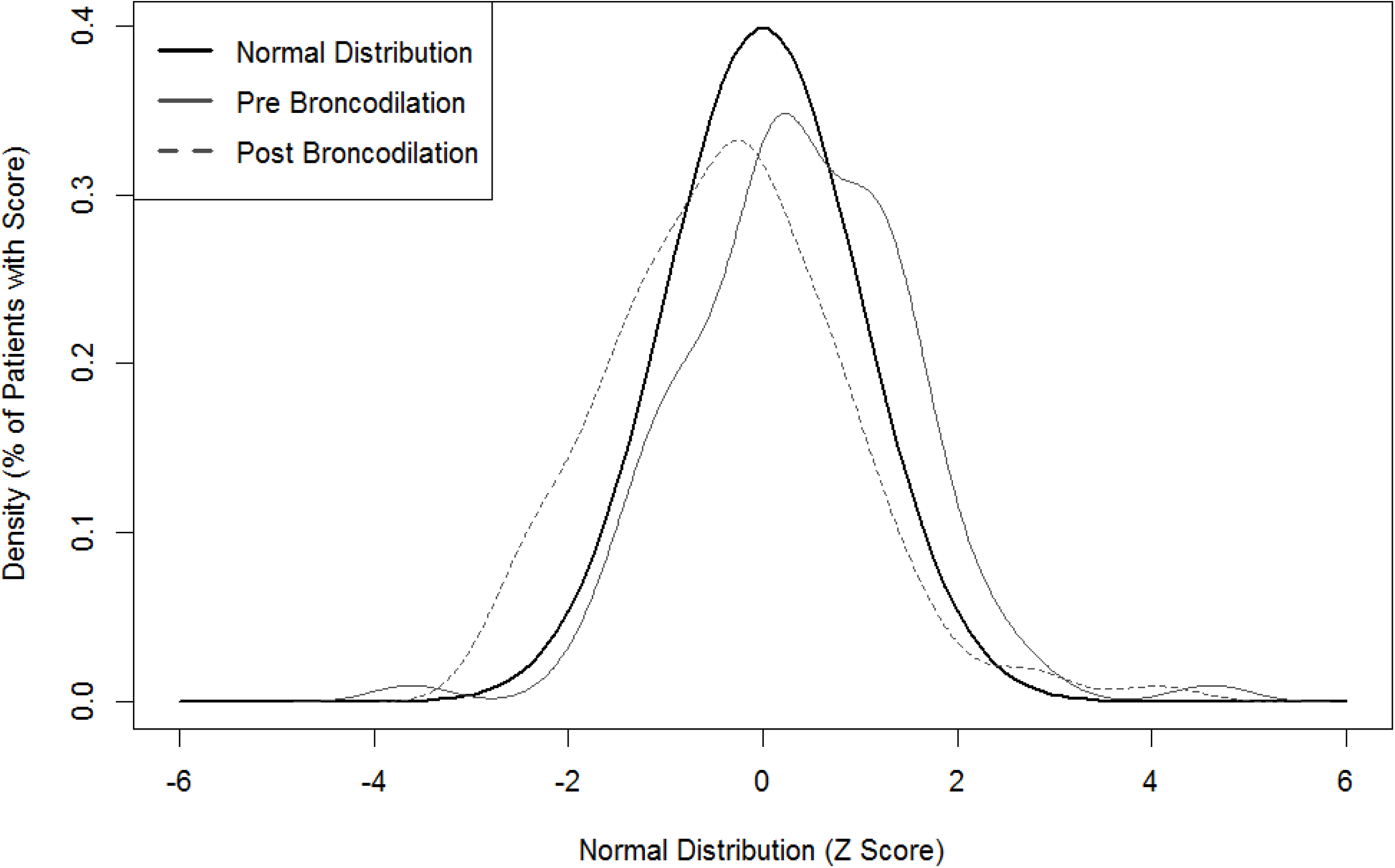
**Distribution of pre- and post-bronchodilator R5 z-scores of healthy adolescents with no history of tuberculosis disease**

**Fig. 2b:**
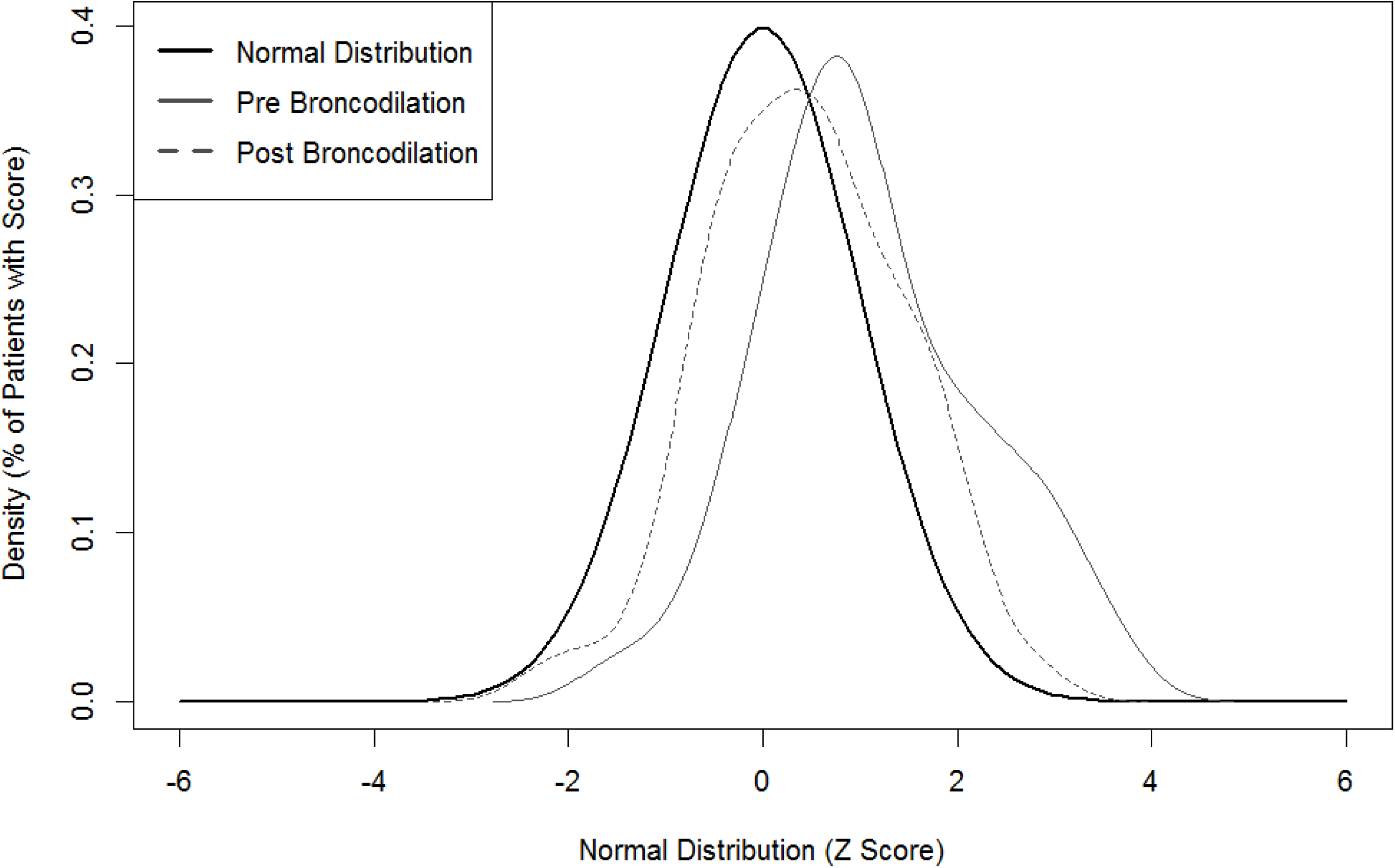
**Distribution of pre- and post-bronchodilator AX z-scores of healthy adolescents with no history of tuberculosis disease**

### Overall assessment of lung health

Lung function measures and both versions of the SGRQ had small correlations with each other, none of which reached statistical significance (Table 2). R5 and AX correlated with one another (0.49, *p* < 0.001) and had negative correlations with FEV1 and FEV1/FVC (-0.38 to -0.31, all with *p* < 0.05). (Lower values are more favorable for oscillometry metrics, whereas higher values are more favorable for spirometry metrics.)

The overall assessment of lung health had strong model fit characteristics using both the original and revised SGRQ. Most differences between the two versions were not significant except the chi square statistic, which was better for the revised version (Table 3).

**Table 3:** Structural Equation Modeling Fit Statistics for the Overall Assessment of Lung Health Using the Revised vs. Original St. George’s Respiratory Questionnaire.

| Model | Chi Square<sup>#</sup> | DF | *p*<sup>#</sup> | CFI | RMSEA | SRMR |
| --- | --- | --- | --- | --- | --- | --- |
| Revised SGRQ, oscillometry, and spirometry | 31.78 | 78 | > 0.99 | 0.993 [0.980, 1.000] | 0.069 [0.022, 0.121] | 0.030 [0.000, 0.093] |
| Original SGRQ, oscillometry, and spirometry | 39.06 | 78 | > 0.99 | 0.990 [0.975, 1.000] | 0.091 [0.053, 0.139] | 0.028 [0.000, 0.112] |
| Difference | 0.87 | 0 | 0.02 | 0.004 [-0.010, 0.019] | -0.023 [-0.021, -0.018] | 0.001 [-0.104, 0.063] |
| Criterion | Smaller is better |  | > 0.05 | > 0.90 | < 0.10 | < 0.10 |

Brackets represent 95% confidence intervals, except for RMSEA, for which the brackets represent the 90% confidence interval, as is conventional due to the characteristic skew for this metric.
<sup>#</sup>The difference statistic for chi square was calculated as a ratio, conceptually similar to how the F statistic is a ratio of chi square distributed random variables. The p-value represents the probability that the chi square ratio is different from 1, implying no difference in model quality.
Abbreviations: CFI, comparative fit index (acceptable values >0.90); CI, confidence interval; DF, degrees of freedom; RMSEA, root mean square error of approximation (acceptable values <0.10); SRMR, standardized root mean square residual (acceptable values <0.10).

## DISCUSSION

PTLD is characterized by diverse clinical characteristics and underlying pathologies that include airway, lung parenchyma, and cardiopulmonary vascular damage.^24^ Because adolescents’ lungs are still developing, TB-associated lung injury during adolescence may result in failure to achieve their expected peak pulmonary function during early adulthood and permanent lung damage, including bronchiectasis.^1,25^ Furthermore, serious respiratory infections, including TB, in early life are associated with the development of chronic obstructive pulmonary disease.^26,27^ Therefore, assessment tools for evaluating PTLD are critical and need to be both feasible and reliable for adolescents in LMICs, which shoulder the greatest TB burden.

Patient-centered care is the first pillar of the World Health Organization’s endTB strategy;^28^ therefore, patient-reported outcomes – including symptoms, activity limitations, and impacts on daily life – should comprise part of the PTLD evaluation. The original SGRQ is lengthy and complicated to score, making it a less feasible tool. We demonstrated high reliability and validity of an abbreviated, easier-to-score version of the SGRQ, suggesting that it provides a good measurement of respiratory disability among adolescent TB survivors in Lima. The questions removed were related to the most severe impacts on daily life and activity restrictions, as well as those addressing activities, such as sex, that not all adolescents engage in. This finding underscores the importance of adapting and validating the SGRQ in adolescents.

In most PTLD research, lung function has been measured using spirometry. Studies published in the last 50 years have demonstrated obstructive, restrictive, and mixed patterns of airflow impairment, as measured by spirometry.^29^ This variability reflects the broad range of lung pathologies that occur with TB, including fibrosis, bronchiectasis, cavity formation, and other architectural distortion.^30^ We observed limitations with spirometry; specifically, not all adolescents could complete the maneuver and that the measurement of FEV1 – and, thus, FEV1/FVC – had suboptimal reliability. Nonetheless, FEV1 and FEV1/FVC had small correlations with total SGRQ score, reflecting the clinical significance of these measures. FVC was acceptably reliable, suggesting that its measurement was less affected by problems using the spirometry device, but it correlated less with SGRQ than FEV1 or FEV1/FVC.

In our cohort, oscillometry was more feasible and reliable, but R5 and AX correlated minimally with the SGRQ. Further research is needed to understand the clinical significance of these abnormal oscillometry metrics. Advanced imaging, such as computed tomography, can clarify underlying anatomical changes. Qualitative research is needed to examine whether the SGRQ – despite our modifications – did not sufficiently capture the effect of lung function abnormalities on adolescents. Finally, longitudinal studies are needed to understand the long-term prognosis for adolescent TB survivors with abnormal oscillometry metrics.

The small or minimal correlations that we observed between the SGRQ, spirometry, and oscillometry – as well as the good model fit of this combination – suggest that these evaluations measure related but distinct aspects of lung health. Thus, our findings support the use of all three tools in the evaluation of adolescent PTLD.

This study had limitations. First, we recorded R5-20 values from oscillometry, which are used to detect small airway disease. However, no reference equations were available to calculate z-scores for this metric. Second, we did not record the number of attempts needed for participants to successful complete the spirometry maneuver, or alternatively, the number of attempts they were able to tolerate before the effort was aborted. This information would have contributed to a more comprehensive understanding of the difficulty of spirometry. Finally, we cannot extrapolate the test characteristics of these tools to adolescent TB survivors in different settings.

In conclusion, our study introduced a shorter, easier-to-score, and validated version of the SGRQ for adolescents; demonstrated feasibility and reliability of spirometry and oscillometry in our cohort; and provided evidence to support use of the Oosteven reference equations and the combination of the SGRQ, spirometry, and oscillometry to evaluate PTLD in adolescent TB survivors. These results pave the way for critical research to better understand the long-term impacts of TB disease on adolescent lung health.

## Data Availability

All data produced in the present study are available upon reasonable request to the authors.

## ACKNOLWEDGEMENTS

This study was funded by the Charles H. Hood Foundation and the National Institutes of Health (R25AI140490). We thank our colleagues at Socios En Salud: Katya León Ostos, Rosa Espinoza Meza, Judith Ramirez Sandoval, Karen Tintaya, Judith Jimenez, and Carmen Capcha. We also thank the Ministry of Health of Peru, including all the front-line health providers who allowed us to recruit and enroll participants at their clinics.

## REFERENCES

1. Chiang SS, Murray MB, Kay AW, Dodd PJ. Factors driving adolescent tuberculosis incidence by age and sex in 30 high-tuberculosis burden countries: a mathematical modelling study. BMJ Glob Health 2025;10(3):e015368. DOI: 10.1136/bmjgh-2024-015368.

2. Dodd PJ, Yuen CM, Jayasooriya SM, van der Zalm MM, Seddon JA. Quantifying the global number of tuberculosis survivors: a modelling study. Lancet Infect Dis 2021;21(7):984–992. DOI: 10.1016/S1473-3099(20)30919-1.

3. Migliori GB, Marx FM, Ambrosino N, et al. Clinical standards for the assessment, management and rehabilitation of post-TB lung disease. Int J Tuberc Lung Dis 2021;25(10):797–813. DOI: 10.5588/ijtld.21.0425.

4. Maleche-Obimbo E, Odhiambo MA, Njeri L, et al. Magnitude and factors associated with post-tuberculosis lung disease in low- and middle-income countries: a systematic review and meta-analysis. PLOS Glob Public Health 2022;2(12):e0000805. DOI: 10.1371/journal.pgph.0000805.

5. van der Zalm MM, Jongen VW, Swanepoel R, et al. Impaired lung function in adolescents with pulmonary tuberculosis during treatment and following treatment completion. EClinMed 2024;67:102406. DOI: 10.1016/j.eclinm.2023.102406.

6. Nkereuwem E, Agbla S, Njai B, et al. Post-tuberculosis respiratory impairment in Gambian children and adolescents: a cross-sectional analysis. Pediatr Pulmonol 2024;59(7):1912–1921. DOI: 10.1002/ppul.27009.

7. Cesilia C, Rinawan FR, Santoso P, Nataprawira HM. Post-TB sequelae in adolescent pulmonary TB survivors. IJTLD Open 2025;2(1):19–25. DOI: 10.5588/ijtldopen.24.0039.

8. Jones PW, Quirk FH, Baveystock CM, Littlejohns P. A self-complete measure of health status for chronic airflow limitation. The St. George’s Respiratory Questionnaire. Am Rev Respir Dis 1992;145(6):1321–7. DOI: 10.1164/ajrccm/145.6.1321.

9. Wilson CB, Jones PW, O’Leary CJ, Cole PJ, Wilson R. Validation of the St. George’s Respiratory Questionnaire in bronchiectasis. Am J Respir Crit Care Med 1997;156(2 Pt 1):536-41. DOI: 10.1164/ajrccm.156.2.9607083.

10. Matheson AM, McIntosh MJ, Kooner HK, et al. Longitudinal follow-up of postacute COVID-19 syndrome: DL(CO), quality-of-life and MRI pulmonary gas-exchange abnormalities. Thorax 2023;78(4):418–421. DOI: 10.1136/thorax-2022-219378.

11. Komarow HD, Myles IA, Uzzaman A, Metcalfe DD. Impulse oscillometry in the evaluation of diseases of the airways in children. Ann Allergy Asthma Immunol 2011;106(3):191–9. DOI: 10.1016/j.anai.2010.11.011.

12. Komarow HD, Skinner J, Young M, et al. A study of the use of impulse oscillometry in the evaluation of children with asthma: analysis of lung parameters, order effect, and utility compared with spirometry. Pediatr Pulmonol 2012;47(1):18–26. DOI: 10.1002/ppul.21507.

13. Maugeri LL, S.; Mellano, E.; Dassetto, D.; Piccioni, P.; Bugiani, M.; Dellaca, R.; Gulotta, C. Force oscillation technique (FOT) vs. spirometry for assessing the imapct of environmental exposure in children. ERS International Congress. Milan, Italy, 2017.

14. Oostveen E, Boda K, van der Grinten CP, et al. Respiratory impedance in healthy subjects: baseline values and bronchodilator response. Eur Respir J 2013;42(6):1513–23. DOI: 10.1183/09031936.00126212.

15. World Health Organization. Global Tuberculosis Report 2024. Geneva, Switzerland: WHO, 2024.

16. Chiang SS, Zeng C, Roman-Sinche B, et al. Adaptation and validation of a TB stigma scale for adolescents in Lima, Peru. Int J Tuberc Lung Dis 2023;27(10):754–760. DOI: 10.5588/ijtld.23.0104.

17. World Health Organization. Definitions and Reporting Framework for Tuberculosis - 2013 Revision. Geneva, Switzerland: WHO, 2013.

18. King GG, Bates J, Berger KI, et al. Technical standards for respiratory oscillometry. Eur Respir J 2020;55(2):1900753. DOI: 10.1183/13993003.00753-2019.

19. Graham BL, Steenbruggen I, Miller MR, et al. Standardization of Spirometry 2019 Update. An Official American Thoracic Society and European Respiratory Society Technical Statement. Am J Respir Crit Care Med 2019;200(8):e70–e88. DOI: 10.1164/rccm.201908-1590ST.

20. Quanjer PH, Stanojevic S, Cole TJ, et al. Multi-ethnic reference values for spirometry for the 3-95-yr age range: the global lung function 2012 equations. Eur Respir J 2012;40(6):1324–43. DOI: 10.1183/09031936.00080312.

21. Ricardo LIC, Wendt A, Costa CDS, et al. Gender inequalities in physical activity among adolescents from 64 Global South countries. J Sport Health Sci 2022;11(4):509–520. DOI: 10.1016/j.jshs.2022.01.007.

22. Raykov T. On structural models for analyzing change. Scand J Psychol 1992;33(3):247–265. (In English). DOI: DOI 10.1111/j.1467-9450.1992.tb00914.x.

23. Harlow L. The essence of multivariate thinking: basic themes and methods. New York, USA: Routledge, 2014.

24. Allwood BW, Byrne A, Meghji J, Rachow A, van der Zalm MM, Schoch OD. Post-tuberculosis lung disease: clinical review of an under-recognised global challenge. Respiration 2021;100(8):751–763. DOI: 10.1159/000512531.

25. Stocks J, Hislop A, Sonnappa S. Early lung development: lifelong effect on respiratory health and disease. Lancet Respir Med 2013;1(9):728–42. DOI: 10.1016/S2213-2600(13)70118-8.

26. Duan P, Wang Y, Lin R, et al. Impact of early life exposures on COPD in adulthood: A systematic review and meta-analysis. Respirology 2021;26(12):1131–1151. DOI: 10.1111/resp.14144.

27. Byrne AL, Marais BJ, Mitnick CD, Lecca L, Marks GB. Risk factors for and origins of COPD. Lancet 2015;385(9979):1723-1724. DOI: 10.1016/S0140-6736(15)60884-4.

28. World Health Organization. The End TB Strategy. Geneva, Switzerland: WHO, 2015. Available at: https://www.who.int/tb/End_TB_brochure.pdf?ua=1. Accessed Sept. 4, 2020.

29. Ivanova O, Hoffmann VS, Lange C, Hoelscher M, Rachow A. Post-tuberculosis lung impairment: systematic review and meta-analysis of spirometry data from 14 621 people. Eur Respir Rev 2023;32(168). DOI: 10.1183/16000617.0221-2022.

30. Byrne A, Al-Hindawi Y, Plit M, et al. The prevalence and pattern of post tuberculosis lung disease including pulmonary hypertension from an Australian TB service; a single-centre, retrospective cohort study. BMC Pulmon Med 2025;25(1):84. DOI: 10.1186/s12890-025-03549-5.

