## Appendix for "Evaluation of New and Repurposed Tools to Assess Post-Tuberculosis Lung Disease in Adolescents: A Cross-Sectional Analysis"

**Number (%) of Response Selections in the St. George's Respiratory Questionnaire among Adolescent Tuberculosis Survivors (N = 101)**

| | | | | | |
|---|---|---|---|---|---|
| **SYMPTOMS SUBSCALE** | | | | | |
| *In the last 3 months, how often have you experienced the following:* | | | | | |
| | *Almost every day of the week* | *Multiple days of the week* | *A few days of the week* | *Only when I have a respiratory infection* | *Not at all* |
| 1. Cough* | 1 (0.99) | 1 (0.99) | 3 (2.97) | 53 (52.58) | 43 (42.57) |
| 2. Bring up phlegm (sputum)* | 0 (0.00) | 0 (0.00) | 5 (4.95) | 39 (38.61) | 57 (56.44) |
| 3. Had difficulty breathing (been short of breath)* | 2 (1.98) | 1 (0.99) | 4 (3.96) | 17 (16.83) | 77 (76.24) |
| 4. Wheezed[#] | 0 (0.00) | 1 (0.99) | 1 (0.99) | 8 (7.92) | 91 (90.10) |
| | *No* | *Yes* | *NA* | | |
| 5. If you have wheezed, is it worse in the morning?[#] | 7 (6.93) | 3 (2.97) | 91 (90.10) | | |
| | *>3* | *3* | *2* | *1* | *0* |
| 6. How many severe attacks of chest problems have you had? | 0 (0.00) | 0 (0.00) | 1 (0.99) | 3 (2.97) | 97 (96.04) |
| | *>1 week* | *≥3 days* | *1-2 days* | *<1 day* | *NA* |
| 7. How long did your worst chest attack last? | 0 (0.00) | 0 (0.00) | 1 (0.99) | 3 (2.97) | 97 (96.04) |
| | *No good days* | *1-2 good days* | *3-4 good days* | *Almost every day is good* | *Every day is good* |
| 8. In the last 3 months, in an average week, how many good days (with few chest problems) have you had?* | 0 (0.00) | 1 (0.99) | 3 (2.97) | 28 (27.72) | 69 (68.32) |
| **ACTIVITIES SUBSCALE** | | | | | |
| *Which activities make you short of breath these days?* | | | | | |
| | *True* | *False* | | | |
| 1. Sitting or lying down | 1 (0.99) | 100 (99.01) | | | |
| 2. Bathing or getting dressed | 1 (0.99) | 100 (99.01) | | | |
| 3. Walking inside the house | 1 (0.99) | 100 (99.01) | | | |
| 4. Walking outside on a flat surface | 5 (4.95) | 96 (95.05) | | | |
| 5. Going up a flight of stairs* | 14 (13.86) | 87 (86.14) | | | |
| 6. Walking up hills* | 31 (30.69) | 70 (69.31) | | | |

| | | | | |
|---|---|---|---|---|
| 7. Playing sports or games* | 22 (21.78) | 79 (78.22) | | |
| *These are questions about how your activities might be affected by your breathing.* | | | | |
| | *True* | *False* | | |
| 8. I take a long time to get washed or dressed | 0 (0.00) | 101 (100.00) | | |
| 9. I cannot take a bath or shower, or I take a long time | 2 (1.98) | 99 (98.02) | | |
| 10. I walk more slowly than other people, or I stop to rest | 5 (4.95) | 96 (95.05) | | |
| 11. Jobs such as housework take a long time, or I have to stop to rest | 4 (3.96) | 97 (96.04) | | |
| 12. If I walk up one flight of stairs, I have to go slowly or stop* | 13 (12.87) | 88 (87.13) | | |
| 13. If I hurry or walk fast, I have to stop or slow down* | 16 (15.84) | 85 (84.16) | | |
| 14. My breathing makes it difficult to do things such as walk up hills, carry things upstairs, light gardening, dance, play bowling, or play golf | 10 (9.90) | 91 (90.10) | | |
| 15. My breathing makes it difficult to do things such as carry heavy loads, dig the garden or shovel snow, jog or walk at 5 miles per hour, play tennis or swim* | 14 (13.86)) | 87 (86.14) | | |
| 16. My breathing makes it difficult to do things such as very heavy manual work, run, cycle, swim fast, or play competitive sports* | 15 (14.85) | 86 (85.15) | | |
| **IMPACT SUBSCALE** | | | | |
| | *My chest problem caused me to stop working/studying* | *My chest problem interferes with my work/studies* | *My chest problem does not affect my work/studies* | *I do not work or study* |
| 1. If you are working or in school, how have your chest problems affected your work/studies (after your TB treatment)? | 1 (0.99) | 3 (2.97) | 97 (96.04) | 0 (0.00) |
| *Some more questions about your cough and breathlessness* | | | | |
| | *True* | *False* | | |
| 2. It hurts me to cough | 4 (3.96) | 97 (96.04) | | |
| 3. My cough makes me tired | 1 (0.99) | 100 (99.01) | | |
| 4. I get breathless when I talk | 0 (0.00) | 101 (100.00) | | |
| 5. I get breathless when I bend over | 1 (0.99) | 100 (99.01) | | |
| 6. My cough or breathing disturbs my sleep[#] | 0 (0.00) | 101 (100.00) | | |
| 7. I get exhausted easily | 0 (0.00) | 101 (100.00) | | |

| | | | |
|---|---|---|---|
| 8. My cough or breathing is embarrassing in public[#] | 1 (0.99) | 100 (99.01) | |
| 9. My chest trouble is a nuisance to my family, friends, or neighbors | 2 (1.98) | 99 (98.02) | |
| 10. I get afraid or panic when I cannot catch my breath | 3 (2.97) | 98 (97.03) | |
| 11. I feel that I am not in control of my chest problem | 1 (0.99) | 100 (99.01) | |
| 12. I do not expect my chest to get any better | 4 (3.96) | 97 (96.04) | |
| 13. I have become frail or an invalid because of my chest | 2 (1.98) | 99 (98.02) | |
| 14. Exercise is not safe for me* | 14 (13.86) | 87 (86.14) | |
| 15. Everything seems too much of an effort | 1 (0.99) | 100 (99.01) | |
| *Questions about your medication* | | | |
| | *True* | *False* | *Not taking medications* |
| 16. My medication does not help me much | 1 (0.99) | 1 (0.99) | 99 (98.02) |
| 17. I get embarrassed using my medication in public | 0 (0.00) | 2 (1.98) | 99 (98.02) |
| 18. I have unpleasant side effects from my medication | 1 (0.99) | 1 (0.99) | 99 (98.02) |
| 19. My medication interferes with my life a lot | 0 (0.00) | 2 (1.98) | 99 (98.02) |
| *We would like to know how your chest problems generally affect your daily life* | | | |
| | *True* | *False* | |
| 20. I can't play sports or games* | 15 (14.85) | 86 (85.15) | |
| 21. I can't go out for fun or recreation | 2 (1.98) | 99 (98.02) | |
| 22. I can't leave the house to go shopping | 0 (0.00) | 101 (100.00) | |
| 23. I can't do housework | 0 (0.00) | 101 (100.00) | |
| 24. I can't move far from bed or a chair | 0 (0.00) | 101 (100.00) | |
| *Here is a list of other activities that your chest problems may keep you from doing* | | | |
| | *True* | *False* | |
| 25. Go out to take a walk or walk the dog | 1 (0.99) | 100 (99.01) | |
| 26. Do housework or gardening | 0 (0.00) | 101 (100.00) | |
| 27. Have sex | 0 (0.00) | 101 (100.00) | |
| 28. Go to church, an internet café, or other place to have fun | 0 (0.00) | 101 (100.00) | |
| 29. Go out when there is bad weather or a lot of smoke | 10 (9.90) | 91 (90.10) | |
| 30. Visit friends or family or play with children | 0 (0.00) | 101 (100.00) | |

| | *The most important problem I have* | *It causes me many problems* | *It causes me some problems* | *It doesn't cause me any problems* |
|---|---|---|---|---|
| 31. How would you describe your lung health?* | 0 (0.00) | 3 (2.97) | 24 (23.76) | 74 (73.27) |
| | *It stops me from doing everything that I like to do* | *It stops me from doing most things I like to do* | *It stops me from doing 1-2 things that I like to do* | *It doesn't stop me from doing anything I like to do* |
| 32. How do your breathing problems affect you?* | 0 (0.00) | 5 (4.95) | 27 (26.73) | 69 (68.32) |

*These questions were retained in the abridged and validated version.
#These questions met the criterion for removal because >90% of participants selected the same answer choice. However, we elected to keep them because of their high clinical relevance.
